# Salience network and cognitive impairment in Parkinson’s disease

**DOI:** 10.1101/2023.10.13.23296825

**Authors:** Brooke E Yeager, Hunter P Twedt, Joel Bruss, Jordan Schultz, Nandakumar S Narayanan

**Author notes:** **Corresponding Author** Nandakumar Narayanan 169 Newton Road, Pappajohn Biomedical Discovery Building—5336 University of Iowa, Iowa City, 52242.

## Abstract

Parkinson’s disease (PD) is a neurodegenerative disease with cognitive as well as motor impairments. While much is known about the brain networks leading to motor impairments in PD, less is known about the brain networks contributing to cognitive impairments. Here, we leveraged resting-state functional magnetic resonance imaging (rs-fMRI) data from the Parkinson’s Progression Marker Initiative (PPMI) to examine network dysfunction in PD patients with cognitive impairment. We tested the hypothesis that cognitive impairments in PD involve altered connectivity of the salience network (SN), a key cortical network that detects and integrates responses to salient stimuli. We used the Montreal Cognitive Assessment (MoCA) as a continuous index of coarse cognitive function in PD. We report two major results. First, in 82 PD patients we found significant relationships between lower intra-network connectivity of the frontoparietal network (FPN; comprising the dorsolateral prefrontal and posterior parietal cortices bilaterally) with lower MoCA scores. Second, we found significant relationships between lower inter-network connectivity between the SN and the basal ganglia network (BGN) and the default mode network (DMN) with lower MoCA scores. These data support our hypothesis about the SN and provide new insights into the brain networks contributing to cognitive impairments in PD.

**Highlights:**

- Cognitive dysfunction is a prominent clinical symptom in Parkinson’s disease
- Functional connectivity of the salience network is important for cognition
- Salience network dynamics are altered in patients with Parkinson’s disease
- Salience network connectivity with other networks is linked to worse cognition in PD

## 1. Introduction

Cognitive impairments of Parkinson’s disease (PD) occur in 30% of newly diagnosed patients (Meireles & Massano, 2012; Narayanan & Albin, 2022) and in 80% of patients within 20 years of disease progression (Foltynie et al., 2004; Hely et al., 2008). The degree of disability from cognitive symptoms in PD can be severe, leading to a diagnosis of mild cognitive impairment or PD dementia (PDD). PD-related cognitive impairments lead to decreased quality of life and increased mortality rates (Lawson et al., 2014; Macleod et al., 2014). Despite the devastating nature of cognitive impairments in PD, there are few effective treatments to address these symptoms because the mechanisms are poorly understood.

Although cognitive impairments in PD can manifest as deficits in nearly all cognitive domains, executive dysfunction is most pronounced (Zgaljardic et al., 2003; Kudlicka et al., 2011). Executive dysfunction involves deficits in inhibition, interference, working memory, cognitive flexibility, and timing (Diamond, 2013; Gilbert & Burgess, 2008; Parker et al., 2013). These high-level cognitive processes are supported by intrinsic functional brain networks, as evidenced by resting-state functional magnetic resonance imaging (rs-fMRI). Specifically, three canonical cortical networks, the 1) default mode network (DMN), 2) frontoparietal network (FPN), and 3) salience network (SN), have a dynamic relationship to support high-level cognition and have been linked to executive function, with the SN playing a role in modulating DMN and FPN activity (Bressler & Menon, 2010; Seeley et al., 2007; Sridharan et al., 2008). In addition, the basal ganglia network (BGN) is of particular significance to PD because dopaminergic deficits in the basal ganglia can profoundly alter functional brain networks (Obeso et al., 2000; Shafiei et al., 2019; Shima et al., 2023). Previous work identified an important link between SN and DMN coupling and cognitive test scores, and also found that striatal-SN connectivity is linked with PD severity (Putcha et al., 2015, 2016). Despite these data, the reliability of SN dysfunction and the SN’s relationship to other brain networks and cognitive impairments in PD are unclear (Badea et al., 2017), and BGN connectivity with cortical canonical networks (DMN, FPN, and SN) to support cognition in PD is unknown.

We tested the hypothesis that cognitive impairments in PD are related altered SN connectivity (Aracil-Bolaños et al., 2019; Putcha et al., 2015, 2016). We took advantage of the Parkinson’s Progression Marker Initiative (PPMI) (Marek et al., 2011), a high-quality database that includes data from a large number of PD patients. We report two main findings: first, we found decreases in FPN intra-network functional connectivity, and second, we found dysfunctional SN-BGN and SN-DMN functional connectivity in PD patients with cognitive impairments. These data implicate the SN as key neural substrate in cognitive decline in PD and could contribute to the discovery of novel biomarkers for cognitive dysfunction in neurodegenerative disease.

## 2. Methods

### 2.1 Study Dataset and Participants

To study the functional network correlates of cognitive impairments in PD, we analyzed resting-state functional magnetic resonance imaging (rs-fMRI) data from patients with genetic and idiopathic PD. We obtained data from the Parkinson’s Progression Marker Initiative (PPMI) database (www.ppmi-info.org/access-data-specimens/download-data; Marek et al., 2011). The PPMI is an open-access data set containing data from over 850 PD patients across 12 countries, providing a comprehensive and externally validated dataset that can be used to readily probe resting-state functional connectivity in PD. The study was approved by the institutional review board of all participating sites. Written informed consent was obtained from all patients before study enrollment. Eighty-three PD participants with rs-fMRI data were included in the present study; data from one participant with noisy rs-fMRI and fewer volume scans were excluded prior to analyses.

### 2.2 Clinical Assessments

The Montreal Cognitive Assessment (MoCA) was used as a continuous index of coarse cognitive function in PD. The MoCA is a widely used and well-validated metric to measure cognition in neurological disease (Dalrymple-Alford et al., 2010; Freitas et al., 2013; Gill et al., 2008; Nasreddine et al., 2005). In our sample of 82 patients with PD, 60 had normal cognition, 20 had mild cognitive impairment, and 2 had moderate cognitive impairment. To index motor symptom severity, we also Motor Unified Parkinson’s Disease Rating Scale (mUPDRS) Part III scores and Hoehn and Yahr staging for each participant to index motor symptom severity. Disease duration was calculated as the time (in months) from the date of initial diagnosis to the date of the participant’s first fMRI session. A comprehensive table of demographic and neuropsychological information can be found in Table 1.

**Table 1.** CONN toolbox network region of interest (ROI) definitions.

| Network name | Seed | *x* | *y* | *z* |
| --- | --- | --- | --- | --- |
| Default mode network (DMN) | Medial prefrontal cortex | 1 | 55 | -3 |
|  | Lateral parietal cortex (L) | -39 | -77 | 33 |
|  | Lateral parietal cortex (R) | 47 | -67 | 29 |
|  | Posterior cingulate cortex | 1 | -61 | 38 |
| Frontoparietal network (FPN) | Lateral prefrontal cortex (L) | -43 | 33 | 28 |
|  | Posterior parietal cortex (L) | -46 | -58 | 49 |
|  | Lateral prefrontal cortex (R) | 41 | 38 | 30 |
|  | Posterior parietal cortex (R) | 52 | -52 | 45 |
| Salience network (SN) | Anterior cingulate cortex | 0 | 22 | 35 |
|  | Anterior insula (L) | -44 | 13 | 1 |
|  | Anterior insula (R) | 47 | 14 | 0 |
|  | Rostral prefrontal cortex (L) | -35 | 45 | 27 |
|  | Rostral prefrontal cortex (R) | 32 | 46 | 27 |
|  | Supramarginal gyrus (L) | -60 | -39 | 31 |
|  | Supramarginal gyrus (R) | 62 | -35 | 32 |

Network ROIs were defined via independent component analysis of Human Connectome Project data (N = 497) (Whitfield-Gabrieli & Nieto-Castanon, 2012). Network seeds are listed with x, y, z coordinates for the centroid of each seed.

### 2.3 MRI Acquisition and Preprocessing

All participants underwent standardized MRI scans on a 3T Siemens Trio Tim scanner at one of nine institutions within the United States and Europe. Full details can be found in the MRI operations manual at http://www.ppmi-info.org/. A 3D T1 image was acquired using the following parameters: repetition time (TR) = 2300 ms, echo time (TE) = 2.98 ms, flip angle (FA) = 9°, and voxel size = 3.3 mm^3^. For rs-fMRI scans, 210 volumes were acquired using the following parameters: TR = 2400 ms, TE = 25 ms, FA = 80°, and voxel size = 3.3 mm^3^. The participants were instructed to rest quietly with their eyes open, clear their mind, and not fall asleep. The rs-fMRI scanning sequence was run for 8 minutes and 24 seconds.

Both T1 structural and rs-fMRI data were downloaded and imported into the functional connectivity (CONN) toolbox (version 21a), which is an open-source MATLAB/SPM-based software (Whitfield-Gabrieli & Nieto-Castanon, 2012). The CONN toolbox default preprocessing pipeline for volume-based analyses was used to preprocess the data, followed by the default denoising pipeline. Briefly, the preprocessing pipeline includes realignment and unwarp for subject motion correction; slice-timing correction; outlier detection with artifact detection tools (ART), which identifies acquisitions with framewise displacement above 1 mm and flags them as outliers; normalization into MNI space; and smoothing with a 6-mm Gaussian kernel. Additionally, anatomical volumes are segmented into gray matter, white matter, and cerebrospinal fluid (CSF). By default, the CONN toolbox does not use global signal regression; however, we chose to remove the effects of undesired global noise and artifact in the analysis of our data by adding a whole-brain mask region of interest (ROI) as an additional confound in our preprocessing pipeline. Next, we implemented the denoising pipeline to remove confounding features using linear regression and applied a temporal band-pass filter (0.01—0.08 Hz), which is typical in rs-fMRI analyses (He et al., 2008; Schölvinck et al., 2010). Estimated confounding subject-motion effects representing three translational and three rotational parameters were removed, as well as five temporal derivatives from the white matter and CSF.

### 2.4 Functional Connectivity Analyses

Seed-based resting state functional connectivity analysis was then performed with the default weighted general linear model used in the CONN toolbox. Seed-to-voxel and region of interest-to-region of interest (ROI-to-ROI) connectivity measures were selected to evaluate ROI-to-ROI functional connectivity. Seed-to-voxel and ROI-to-ROI Pearson’s correlation connectivity maps for each participant were computed in CONN. First-level correlations were Fisher r-to-z transformed and exported as subject-level z-maps to improve normality assumptions of our models. Second-level analyses were computed with MATLAB (R2022b) and R (version 4.3.1) to make inferences about group differences. To find intra-network connectivity of each network, z-values of each ROI-to-ROI pair within a single network were averaged. To find inter-network connectivity, Fisher-transformed values of each ROI-to-ROI pair for two given networks were averaged.

Cortical and subcortical ROIs were derived from the Harvard-Oxford atlas, while cerebellar ROIs were identified from the automated anatomical labeling (AAL) atlas. The basal ganglia network (BGN) was calculated using 12 ROIs from the Harvard-Oxford atlas: left and right caudate; left and right putamen; left and right pallidum; left and right hippocampus; left and right amygdala; and left and right accumbens. These ROIs were selected to calculate BGN functional connectivity based on previous work (Luo et al., 2012). The network ROIs used by the CONN toolbox were derived from independent component analysis of the Human Connectome Project dataset (N = 497) (Whitfield-Gabrieli & Nieto-Castanon, 2012). Three cognitive networks found in the literature on resting-state brain network (default mode network (DMN); frontoparietal network (FPN); and salience network (SN) (Aracil-Bolaños et al., 2019; Baggio et al., 2014, 2015; Chen et al., 2022; Goulden et al., 2014; Lewis et al., 2012; Lucas-Jiménez et al., 2016; Putcha et al., 2015, 2016)) were selected for further analysis based on our *a priori* hypotheses that SN connectivity within and between these networks support cognition and are impaired in PD patients with cognitive impairment. The composition of each network is further defined in Table 1.

### 2.5 Statistical Analyses

We constructed linear regression models to examine functional connectivity relationships with MoCA scores. For qualitative network comparisons, participants with MoCA scores greater than or equal to 26 were classified as being cognitively normal (PD-Norm), whereas those with MoCA scores less than or equal to 25 were classified as having cognitive impairment (PD-CI).

All statistical relationships among clinical assessments, demographic data, and connectivity were calculated via Spearman’s correlation. Variables that had significant univariate relationships with MoCA scores (Table 2) were included as covariates in our models (*lm* in R; MoCA ∼ FC + Disease Duration + Age + Education). Effect sizes were calculated via partial eta^2^ (*etaSquared* in R). We interpreted p-values of 0.05 or less as significant. R (version 4.3.1) was used for all analyses. All data and code are available at narayanan.lab.uiowa.edu.

**Table 2.**
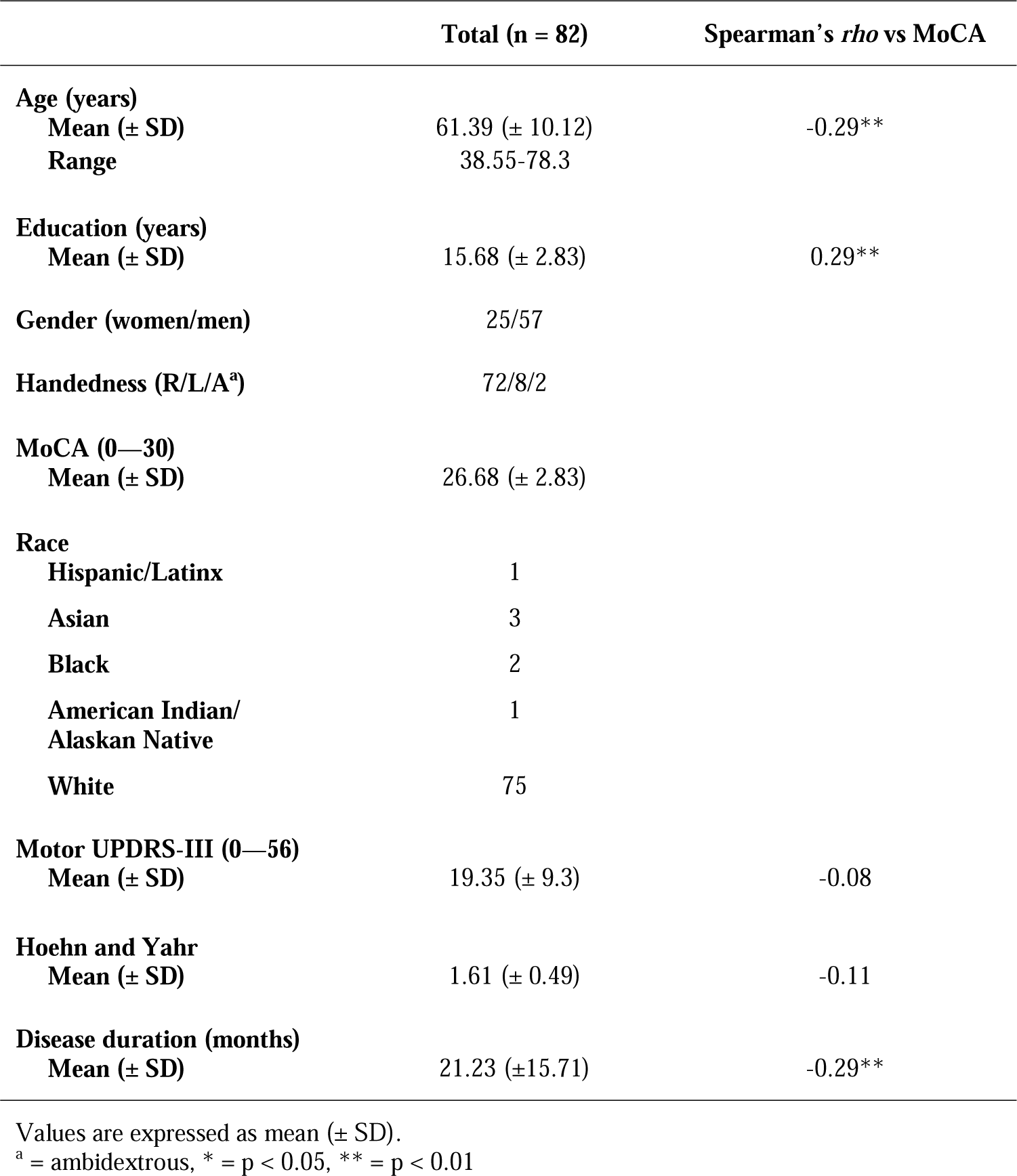
Demographic and clinical data of study population.

## 3. Results

Demographic data from our sample of 82 PD patients from the PPMI database are described in Table 2. Spearman’s correlations revealed significant relationships for MoCA and disease duration, age, and education (Table 2). We included variables that had a strong univariate relationship with MoCA in multivariate models of connectivity (Anjum et al., 2023; Dalrymple-Alford et al., 2010; Freitas et al., 2013; Gill et al., 2008; Hendershott et al., 2017; Kandiah et al., 2014; Litvan et al., 2012; Nasreddine et al., 2005; Singh et al., 2021; Zadikoff et al., 2008).

### 3.1 Intra-network Connectivity

We investigated the relationship between functional network connectivity and cognition in PD with a focus on the SN. First, we examined intra-network connectivity of the SN as a function of cognitive status as defined by the MoCA. While generally used as a screening tool for cognitive impairments in PD, the MoCA is widely used, can sensitively detect cognitive impairments and is comparable across other studies (Cole et al., 2023; Dalrymple-Alford et al., 2010; Singh et al., 2018, 2021). Contrary to our hypothesis, we did not find a significant relationship between intra-SN functional connectivity and cognition (*p* = 0.8), but we did find a significant relationship with age (*p* = 0.04).

We examined intra-network connectivity of DMN, FPN, and BGN as a function of cognitive status as defined by the MoCA. When controlling for significant univariate predictors of MoCA scores (Table 2), we found a significant relationship of intra-FPN functional connectivity as a function of MOCA (*β* = 3.53, *p* = 0.01, *eta^2^* = 0.08; Fig 1A—C). Of note, we did not observe reliable relationships for FPN with the mUPDRS Part III scores (*r* = −0.06, *p* = 0.57). We also did not find reliable relationships of intra-DMN or BGN connectivity and cognition.

**Figure 1.**
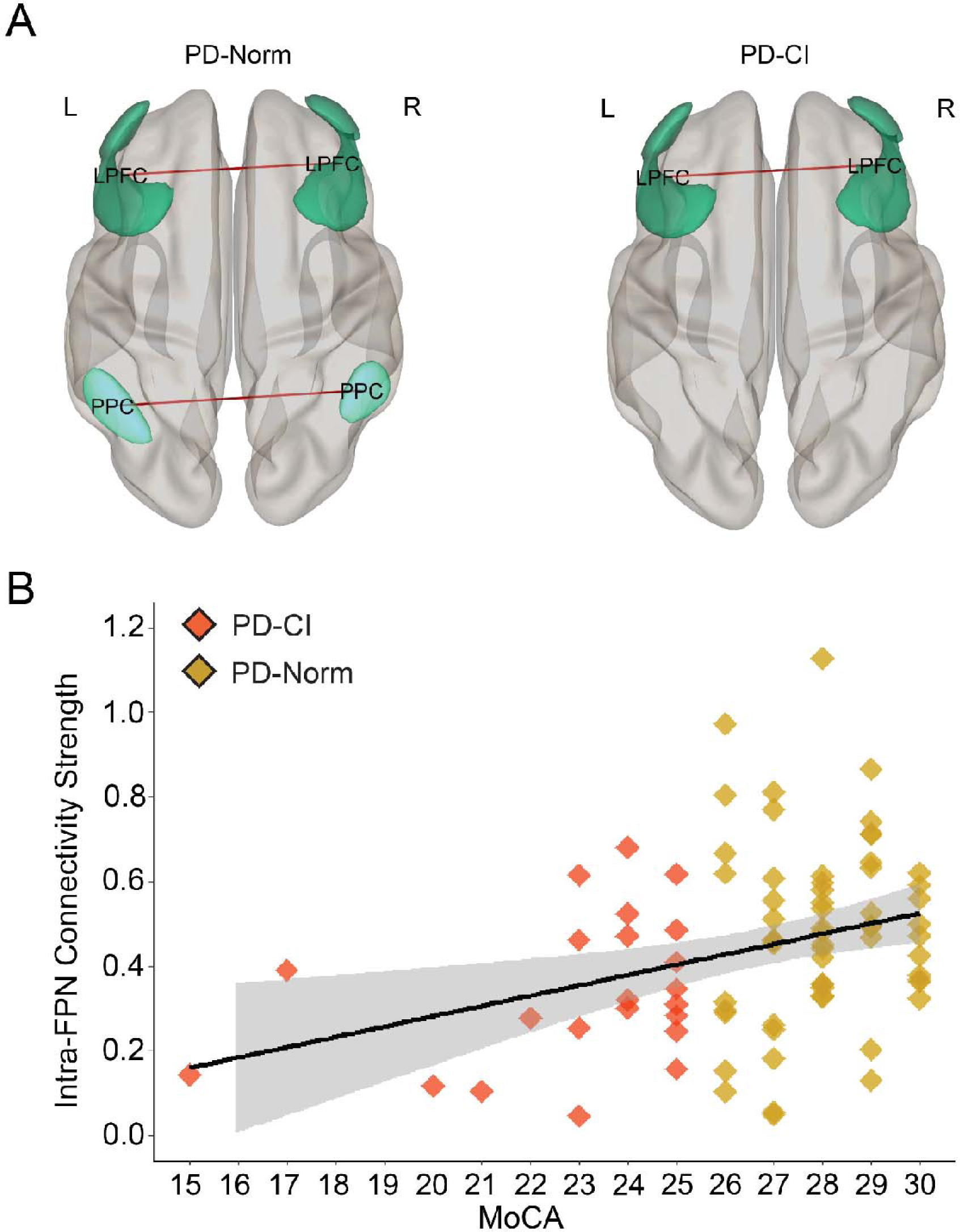
Intra-FPN functional connectivity. A) 3D rendered display of supra-threshold (p < 0.05) ROI-level results for intra-FPN functional connectivity shown for PD patients with normal cognition (PD-Norm; *left*) and PD patients with cognitive impairments (PD-CI; *right*). Red line indicate positive associations; line width is proportional to degree of connectivity. B) Scatterplot displaying a significant relationship between intra-FPN functional connectivity (Fisher r-to-z values) and MoCA scores. Gray band = 95% confidence interval. *LPFC = lateral prefrontal cortex, PPC = posterior parietal cortex*.

### 3.2 Inter-network Connectivity

Next, we examined inter-network functional connectivity between SN, DMN, FPN and BGN networks. Again, when controlling for significant univariate predictors of MoCA scores (Table 2), we found that more positive SN-BGN functional connectivity was associated with higher MoCA scores (*β* = 8.46, *p* = 0.02; *eta^2^* = 0.07; Fig 2A—C). We also found a significant relationship between more positive SN-DMN functional connectivity and higher MoCA scores (*β* = 4.83, *p* = 0.04; *eta^2^* = 0.05; Fig 3A—C). These data supported the idea that SN network connectivity contributes to cognitive impairments in PD (Putcha et al., 2015, 2016). SN-BGN and SN-DMN functional connectivity were not related to mUPDRS Part III scores (*r* = −0.05, *p* = 0.65; *r* = −0.08, *p* = 0.49). We did not find reliable relationships between SN-FPN, DMN-FPN, DMN-BGN or FPN-BGN connectivity and MoCA. Together, these data further implicate the importance of SN network connectivity in cognitive impairments in PD.

**Figure 2.**
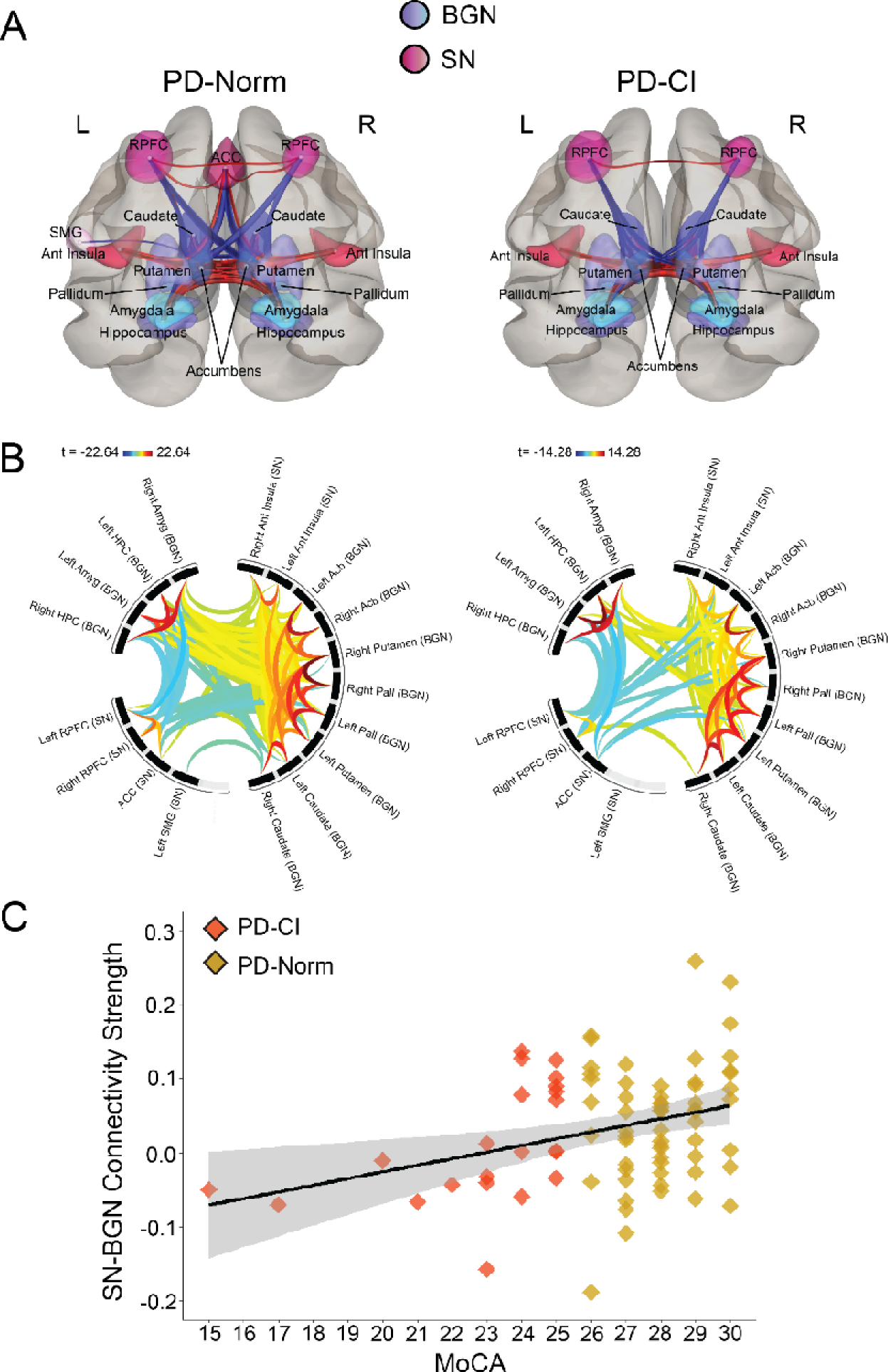
SN-BGN functional connectivity. A) 3D rendered display of supra-threshold (p < 0.05) ROI-level results for SN-BGN functional connectivity shown for PD patients with normal cognition (PD-Norm; *left*) and PD patients with cognitive impairments (PD-CI; *right*). Red line indicate positive associations, and blue lines indicate negative associations; line width is proportional to degree of connectivity. B) Connectome ring display with significant clusters of SN-BGN connections. Results were corrected for multiple comparisons using false discovery rate (FDR) across all possible pairwise clusters. Color bar represents statistical t value where warm colors represent positive correlations and cooler colors represent negative correlations. C) Scatterplot displaying a significant relationship between SN-BGN functional connectivity (Fisher r-to-z values) and MoCA scores. Gray band = 95% confidence interval. *RPFC = rostral prefrontal cortex, SMG = supramarginal gyrus, Ant Insula = anterior insula, ACC = anterior cingulate cortex, Amyg = amygdala, HPC = hippocampus, Pall = pallidum, Acb = accumbens*

**Figure 3.**
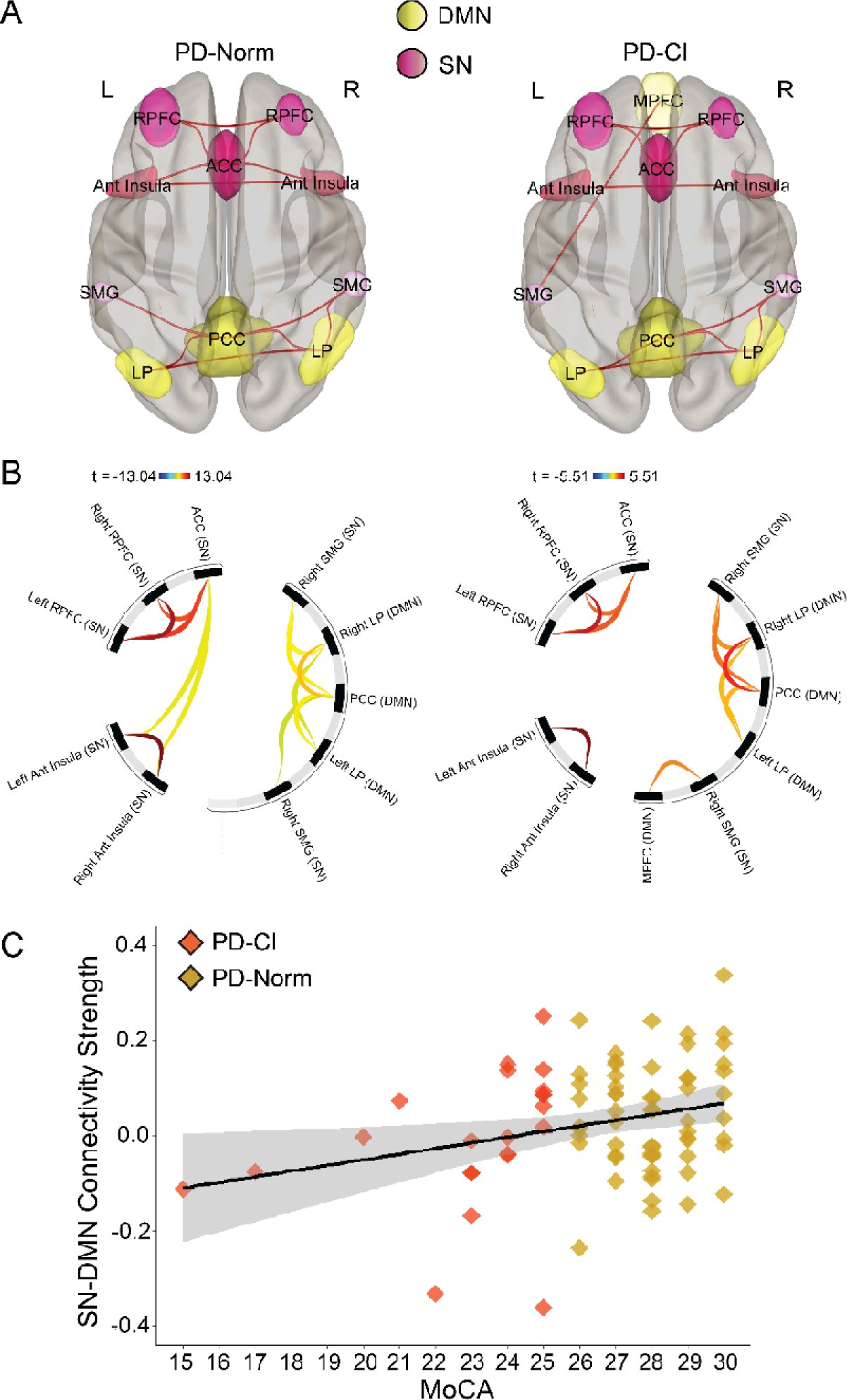
SN-DMN functional connectivity. A) 3D rendered display of supra-threshold (p < 0.05) ROI-level results for SN-DMN functional connectivity shown for PD patients with normal cognition (PD-Norm; *left*) and PD patients with cognitive impairments (PD-CI; *right*). Red line indicate positive associations, and blue lines indicate negative associations; line width is proportional to degree of connectivity. B) Connectome ring display with significant clusters of SN-DMN connections. Results were corrected for multiple comparison using false discovery rat (FDR) across all possible pairwise clusters. Color bar represents statistical t value where warm colors represent positive correlations and cooler colors represent negative correlations. C) Scatterplot displaying a significant relationship between SN-DMN functional connectivity (Fisher r-to-z values) and MoCA scores. Gray band = 95% confidence interval. *MPFC = medial prefrontal cortex, PCC = posterior cingulate cortex, LP = lateral parietal.*

## 4. Discussion

We tested the hypothesis that cognitive impairments in PD are associated with decreased SN connectivity. In our sample of 82 PPMI patients with PD, we found that lower MoCA scores had decreased intra-FPN functional connectivity and decreased inter-SN-BGN and SN-DMN functional connectivity. Our results provide new information to improve our understanding of the brain networks contributing to cognitive impairments in PD.

In line with previous work, we show reduced intra-FPN functional connectivity with worse cognition (Amboni et al., 2015). Specifically, our results point to alterations in posterior parietal nodes of the FPN in PD-CI patients (Fig 1A). It is possible that these posterior parietal connectivity alterations are driven by the metabolic abnormalities in the parietal cortex that are hallmarks in PD (Firbank et al., 2017; Isaias et al., 2020). Reduced parietal glucose metabolism has been linked to cognitive impairments in PD, further supporting this notion (Huang et al., 2007).

Putcha et al. (2015, 2016) and Aracil-Bolaños et al. (2018) found reduced SN-DMN functional connectivity with cognitive impairments in PD which we replicate in the current study. We also extend this work to corticostriatal relationships by showing that SN-BGN functional connectivity predicts cognitive impairments as measured by the MoCA. Putcha et al. (2015) examined functional connectivity with a striatal ROI consisting of bilateral caudate and putamen; we expand on these results by including an entire BGN ROI which includes additional cortical and subcortical structures affected in PD. Unlike previous work, we did not find evidence of altered SN-FPN or DMN-FPN connectivity predicting cognitive impairments (Amboni et al., 2015; Putcha et al., 2015). Together, our results support and expand upon previous work finding important relationships between intra-FPN, SN-BGN, SN-DMN functional connectivity and cognition in PD (Aracil-Bolaños et al., 2019; Putcha et al., 2015, 2016).

The SN is consistently composed of the midcingulate cortex and insula, and has been implicated in detecting and integrating responses to salient stimuli (Menon & Uddin, 2010; Seeley et al., 2007). The SN may influence the dynamic relationship between the DMN and FPN (Goulden et al., 2014; Menon, 2011; Sridharan et al., 2008) and help switch between the DMN and FPN during goal-directed behaviors. Control by the SN over the DMN and FPN is dysregulated in several psychiatric and neurological disorders (Chand et al., 2017; Menon, 2011; Seeley et al., 2007), including PD (Putcha et al., 2015, 2016). Our work here adds to this evidence linking SN functional connectivity with other large-scale brain networks and cognitive impairments in PD. Specifically, we show that more positive functional connectivity between the SN-DMN is linked to better cognition in PD which may suggest that positive coupling between these networks is necessary for the SN to efficiently disengage the DMN during cognitive control.

The regions making up the SN can be affected by PD-relevant pathological processing (Vogel et al., 2023). Marked gray matter atrophy and reduced cerebral blood flow can be seen in the anterior cingulate cortex (ACC) (Lewis et al., 2012; Nagano-Saito et al., 2005; Summerfield et al., 2005), a key node of the SN (Uddin, 2016). The cingulate cortex and amygdala are also directly affected by synucleinopathy; in a post-mortem sample of 53 patients with PD, alpha-synuclein inclusions were found in the cingulate cortex in 34% of cases and in the amygdala in 24% of cases (Jellinger, 2003). Moreover, degradation of dopaminergic afferents to the ACC is characteristic of PD, and dopamine plays an essential role in high-level cognition (Alberico et al., 2015; Ko et al., 2009; Lumme et al., 2007; Vogt, 2019). Dopamine also directly impacts large-scale network architecture. Depleting dopamine increases signal variability of the SN which in turn makes synchronizing neuronal populations more difficult, leading to decreased corticostriatal connectivity (Shafiei et al., 2019; Shima et al., 2023). In support of this idea, we found altered SN-BGN functional connectivity associated with cognitive impairments in PD.

Our results with rs-fMRI implicating the SN as a key neural substrate of cognitive impairments in PD is consistent with previous human electroencephalography (EEG) work. High level cognition, such as cognitive control, is supported by low-frequency neural activity over mid-frontal brain areas, and this signal is thought to be at least partially generated by the cingulate cortex (Cavanagh & Frank, 2014). In patients with PD, there is an attenuation of low-frequency mid-frontal neural activity and this is associated with cognitive dysfunction (Cole et al., 2023; Narayanan et al., 2013; Parker et al., 2015; Singh et al., 2018, 2021, 2023; Uc et al., 2023). These rhythms are trigged by task-relevant cues that engage SN networks, which may in turn engage the BGN to coordinate a cognitively controlled response.

Our work has several limitations. First, unlike previous literature (Baggio et al., 2015; Chen et al., 2022; de la Cruz et al., 2020; Lucas-Jiménez et al., 2016; Tessitore et al., 2012, 2019), we did not find any relationships between DMN functional connectivity and cognition in PD, which may be attributed to the vast heterogeneity of PD (Badea et al., 2017). Second, in our sample of patients with PD, we only include two patients who have MoCA scores low enough to be considered PDD. Although challenging, this is key for investigating neural circuitry associated with cognitive decline in PD. Third, the MoCA is a measure of coarse global cognitive function, and it is sensitive and robust in detecting cognitive impairments in PD (Dalrymple-Alford et al., 2010; Litvan et al., 2012). Two advantages of the MoCA are 1) it is a highly-used and widely accessible screening tool, and 2) it has a wider scoring range (15—30 in our sample), enabling more detail than simply stratifying patients into PD, PD-MCI, and PDD. The MoCA is also related to traditional cognitive tests of executive function, some of which are not available in all patients in the PPMI database. We cannot exclude, however, that some of our correlations were driven by the particular MoCA distribution in this study. Future work will include specific tests of cognition to further expound upon the relationship between functional connectivity and cognitive impairments in PD. Fourth, the SN defined in the current study included the left and right rostral prefrontal cortex (RPFC). While several studies have included the RPFC as a node of the SN (Almdahl et al., 2023; Cermakova et al., 2023; Tikàsz et al., 2020; Ueno et al., 2020; Webb et al., 2021), this is still in contrast to other studies that have used different parcellations of the SN (Aracil-Bolaños et al., 2019; Dosenbach et al., 2008; S. Marek & Dosenbach, 2018; Menon & Uddin, 2010; Putcha et al., 2015, 2016; Seeley et al., 2007). Finally, the BGN is a complex network, and the structures that make up this network, such as the striatum, have distinct subdivisions with differential patterns of functional connectivity (Di Martino et al., 2008). The BGN defined and calculated in the current study does not consider these distinct subdivisions, and it is possible that potentially important independent signals are being averaged out. Thus, future studies will employ a finer parcellation of the BGN to better parse out the differential patterns of BGN functional connectivity with the SN and other cognitive networks.

## 5. Conclusions

Our current study provides new insight into network dysfunction of cognitive impairments in PD. We find that intra-FPN functional connectivity is linked to cognition, such that lower intrinsic connectivity is seen in patients with worse cognition. Furthermore, we find that a specific relationship of disrupted inter-SN functional connectivity with the BGN and DMN is linked to worse cognition. Our work illuminates the SN as a key network implicated in cognitive impairments in PD. This work could inspire novel biomarkers for cognitive dysfunction in PD and in other neurodegenerative diseases.

## Contributors

BEY conceptualized and designed the study. BEY, HPT, and JB conducted the analyses. BEY, HPT, JB, JS, and NSN interpreted the data. BEY wrote the original draft of the manuscript. BEY, HPT, JB, JS, and NSN edited and revised the manuscript.

## Ethics approval

The PPMI study was approved by the local Institutional Review Boards of respective institutions (a full list is available at the following link https://www.ppmi-info.org/about-ppmi/ppmi-clinical-sites). Written informed consent were obtained from each participant at enrollment, in accordance with the Declaration of Helsinki. All methods were performed in accordance with the relevant guidelines and regulations.

## Data availability statement

Data are available upon reasonable request. All data and code are available at narayanan.lab.uiowa.edu.

## Data Availability

https://www.ppmi-info.org/access-dataspecimens/download-data

## Acknowledgments

Data used in the preparation of this article were obtained June 25, 2021 from the Parkinson’s Progression Markers Initiative (PPMI) database (www.ppmi-info.org/access-dataspecimens/download-data), RRID:SCR 006431. For up-to-date information on the study, visit www.ppmi-info.org. PPMI – a public-private partnership – is funded by the Michael J. Fox Foundation for Parkinson’s Research and funding partners (see full funding partner list at https://www.ppmi-info.org/about-ppmi/who-we-are/study-sponsors).

We would also like to thank Patrick Ten Eyck and Linder H. Wendt for their statistical expertise and advise, as well as Heather A. Widmayer for providing careful and constructive editorial feedback.

## References

Alberico, S. L., Cassell, M. D., & Narayanan, N. S. (2015). The vulnerable ventral tegmental area in Parkinson’s disease. Basal Ganglia, 5(2), 51–55. 10.1016/j.baga.2015.06.001

Almdahl, I. S., Martinussen, L. J., Ousdal, O. T., Kraus, M., Sowa, P., Agartz, I., & Korsnes, M. S. (2023). Task-based functional connectivity reveals aberrance with the salience network during emotional interference in late-life depression. Aging & Mental Health, 1–9. 10.1080/13607863.2023.2179972

Amboni, M., Tessitore, A., Esposito, F., Santangelo, G., Picillo, M., Vitale, C., Giordano, A., Erro, R., de Micco, R., Corbo, D., Tedeschi, G., & Barone, P. (2015). Resting-state functional connectivity associated with mild cognitive impairment in Parkinson’s disease. Journal of Neurology, 262(2), 425–434. 10.1007/s00415-014-7591-5

Anjum, M. F., Espinoza, A., Cole, R., Singh, A., May, P., Uc, E., Dasgupta, S., & Narayanan, N. (2023). *Resting-state EEG measures cognitive impairment in Parkinson’s disease* [Preprint]. In Review. 10.21203/rs.3.rs-2666578/v1

Aracil-Bolaños, I., Sampedro, F., Marín-Lahoz, J., Horta-Barba, A., Martínez-Horta, S., Botí, M., Pérez-Pérez, J., Bejr-Kasem, H., Pascual-Sedano, B., Campolongo, A., Izquierdo, C., Gironell, A., Gómez-Ansón, B., Kulisevsky, J., & Pagonabarraga, J. (2019). A divergent breakdown of neurocognitive networks in Parkinson’s Disease mild cognitive impairment. Human Brain Mapping, 40(11), 3233–3242. 10.1002/hbm.24593

Badea, L., Onu, M., Wu, T., Roceanu, A., & Bajenaru, O. (2017). Exploring the reproducibility of functional connectivity alterations in Parkinson’s disease. PLOS ONE, 12(11), e0188196. 10.1371/journal.pone.0188196

Baggio, H.-C., Sala-Llonch, R., Segura, B., Marti, M.-J., Valldeoriola, F., Compta, Y., Tolosa, E., & Junqué, C. (2014). Functional brain networks and cognitive deficits in Parkinson’s disease. Human Brain Mapping, 35(9), 4620–4634. 10.1002/hbm.22499

Baggio, H.-C., Segura, B., Sala-Llonch, R., Marti, M.-J., Valldeoriola, F., Compta, Y., Tolosa, E., & Junqué, C. (2015). Cognitive impairment and resting-state network connectivity in Parkinson’s disease. Human Brain Mapping, 36(1), 199–212. 10.1002/hbm.22622

Bressler, S. L., & Menon, V. (2010). Large-scale brain networks in cognition: Emerging methods and principles. Trends in Cognitive Sciences, 14(6), 277–290. 10.1016/j.tics.2010.04.004

Cavanagh, J. F., & Frank, M. J. (2014). Frontal theta as a mechanism for cognitive control. Trends in Cognitive Sciences, 18(8), 414–421. 10.1016/j.tics.2014.04.012

Cermakova, P., Chlapečka, A., Csajbók, Z., Andrýsková, L., Brázdil, M., & Marečková, K. (2023). Parental education, cognition and functional connectivity of the salience network. Scientific Reports, 13(1), Article 1. 10.1038/s41598-023-29508-w

Chand, G. B., Wu, J., Hajjar, I., & Qiu, D. (2017). Interactions of the Salience Network and Its Subsystems with the Default-Mode and the Central-Executive Networks in Normal Aging and Mild Cognitive Impairment. Brain Connectivity, 7(7), 401–412. 10.1089/brain.2017.0509

Chen, L., Huang, T., Ma, D., & Chen, Y.-C. (2022). Altered Default Mode Network Functional Connectivity in Parkinson’s Disease: A Resting-State Functional Magnetic Resonance Imaging Study. Frontiers in Neuroscience, 16. https://www.frontiersin.org/articles/10.3389/fnins.2022.905121

Cole, R. C., Espinoza, A. I., Singh, A., Berger, J. I., Cavanagh, J. F., Wessel, J. R., Greenlee, J. D., & Narayanan, N. S. (2023). Novelty-induced frontal–STN networks in Parkinson’s disease. Cerebral Cortex, 33(2), 469–485. 10.1093/cercor/bhac078

Dalrymple-Alford, J. C., MacAskill, M. R., Nakas, C. T., Livingston, L., Graham, C., Crucian, G. P., Melzer, T. R., Kirwan, J., Keenan, R., Wells, S., Porter, R. J., Watts, R., & Anderson, T. J. (2010). The MoCA: Well-suited screen for cognitive impairment in Parkinson disease. Neurology, 75(19), 1717–1725. 10.1212/WNL.0b013e3181fc29c9

de la Cruz, F., Wagner, G., Schumann, A., Suttkus, S., Güllmar, D., Reichenbach, J. R., & Bär, K. (2020). Interrelations between dopamine and serotonin producing sites and regions of the default mode network. Human Brain Mapping, 42(3), 811–823. 10.1002/hbm.25264

Di Martino, A., Scheres, A., Margulies, D. S., Kelly, A. M. C., Uddin, L. Q., Shehzad, Z., Biswal, B., Walters, J. R., Castellanos, F. X., & Milham, M. P. (2008). Functional Connectivity of Human Striatum: A Resting State fMRI Study. Cerebral Cortex, 18(12), 2735–2747. 10.1093/cercor/bhn041

Diamond, A. (2013). Executive Functions. Annual Review of Psychology, 64(1), 135–168. 10.1146/annurev-psych-113011-143750

Dosenbach, N. U. F., Fair, D. A., Cohen, A. L., Schlaggar, B. L., & Petersen, S. E. (2008). A dual-networks architecture of top-down control. Trends in Cognitive Sciences, 12(3), 99–105. 10.1016/j.tics.2008.01.001

Firbank, M. J., Yarnall, A. J., Lawson, R. A., Duncan, G. W., Khoo, T. K., Petrides, G. S., O’Brien, J. T., Barker, R. A., Maxwell, R. J., Brooks, D. J., & Burn, D. J. (2017). Cerebral glucose metabolism and cognition in newly diagnosed Parkinson’s disease: ICICLE-PD study. *Journal of Neurology*, Neurosurgery & Psychiatry, 88(4), 310–316. 10.1136/jnnp-2016-313918

Foltynie, T., Brayne, C. E. G., Robbins, T. W., & Barker, R. A. (2004). The cognitive ability of an incident cohort of Parkinson’s patients in the UK. The CamPaIGN study. Brain, 127(3), 550–560. 10.1093/brain/awh067

Freitas, S., Simões, M. R., Alves, L., & Santana, I. (2013). Montreal Cognitive Assessment: Validation Study for Mild Cognitive Impairment and Alzheimer Disease. Alzheimer Disease & Associated Disorders, 27(1), 37. 10.1097/WAD.0b013e3182420bfe

Gilbert, S. J., & Burgess, P. W. (2008). Executive function. Current Biology, 18(3), R110–R114. 10.1016/j.cub.2007.12.014

Gill, D. J., Freshman, A., Blender, J. A., & Ravina, B. (2008). The montreal cognitive assessment as a screening tool for cognitive impairment in Parkinson’s disease. Movement Disorders, 23(7), 1043–1046. 10.1002/mds.22017

Goulden, N., Khusnulina, A., Davis, N. J., Bracewell, R. M., Bokde, A. L., McNulty, J. P., & Mullins, P. G. (2014). The salience network is responsible for switching between the default mode network and the central executive network: Replication from DCM. NeuroImage, 99, 180–190. 10.1016/j.neuroimage.2014.05.052

He, B. J., Snyder, A. Z., Zempel, J. M., Smyth, M. D., & Raichle, M. E. (2008). Electrophysiological correlates of the brain’s intrinsic large-scale functional architecture. Proceedings of the National Academy of Sciences, 105(41), 16039–16044. 10.1073/pnas.0807010105

Hely, M. A., Reid, W. G. J., Adena, M. A., Halliday, G. M., & Morris, J. G. L. (2008). The Sydney multicenter study of Parkinson’s disease: The inevitability of dementia at 20 years. Movement Disorders, 23(6), 837–844. 10.1002/mds.21956

Hendershott, T. R., Zhu, D., Llanes, S., & Poston, K. L. (2017). Domain-specific accuracy of the Montreal Cognitive Assessment subsections in Parkinson’s disease. Parkinsonism & Related Disorders, 38, 31–34. 10.1016/j.parkreldis.2017.02.008

Huang, C., Mattis, P., Tang, C., Perrine, K., Carbon, M., & Eidelberg, D. (2007). Metabolic brain networks associated with cognitive function in Parkinson’s disease. NeuroImage, 34(2), 714–723. 10.1016/j.neuroimage.2006.09.003

Isaias, I. U., Brumberg, J., Pozzi, N. G., Palmisano, C., Canessa, A., Marotta, G., Volkmann, J., & Pezzoli, G. (2020). Brain metabolic alterations herald falls in patients with Parkinson’s disease. Annals of Clinical and Translational Neurology, 7(4), 579–583. 10.1002/acn3.51013

Jellinger, K. A. (2003). α-Synuclein pathology in Parkinson’s and Alzheimer’s disease brain: Incidence and topographic distribution—a pilot study. Acta Neuropathologica, 106(3), 191–202. 10.1007/s00401-003-0725-y

Kandiah, N., Zhang, A., Cenina, A. R., Au, W. L., Nadkarni, N., & Tan, L. C. (2014). Montreal Cognitive Assessment for the screening and prediction of cognitive decline in early Parkinson’s disease. Parkinsonism & Related Disorders, 20(11), 1145–1148. 10.1016/j.parkreldis.2014.08.002

Ko, J. H., Ptito, A., Monchi, O., Cho, S. S., Van Eimeren, T., Pellecchia, G., Ballanger, B., Rusjan, P., Houle, S., & Strafella, A. P. (2009). Increased dopamine release in the right anterior cingulate cortex during the performance of a sorting task: A [11C]FLB 457 PET study. NeuroImage, 46(2), 516–521. 10.1016/j.neuroimage.2009.02.031

Kudlicka, A., Clare, L., & Hindle, J. V. (2011). Executive functions in Parkinson’s disease: Systematic review and meta-analysis. Movement Disorders, 26(13), 2305–2315. 10.1002/mds.23868

Lawson, R. A., Yarnall, A. J., Duncan, G. W., Khoo, T. K., Breen, D. P., Barker, R. A., Collerton, D., Taylor, J.-P., & Burn, D. J. (2014). Severity of mild cognitive impairment in early Parkinson’s disease contributes to poorer quality of life. Parkinsonism & Related Disorders, 20(10), 1071–1075. 10.1016/j.parkreldis.2014.07.004

Lewis, S. J. G., Shine, J. M., Duffy, S., Halliday, G., & Naismith, S. L. (2012). Anterior cingulate integrity: Executive and neuropsychiatric features in Parkinson’s disease. Movement Disorders, 27(10), 1262–1267. 10.1002/mds.25104

Litvan, I., Goldman, J. G., Tröster, A. I., Schmand, B. A., Weintraub, D., Petersen, R. C., Mollenhauer, B., Adler, C. H., Marder, K., & Williams-Gray, C. H. (2012). Diagnostic criteria for mild cognitive impairment in Parkinson’s disease: Movement Disorder Society Task Force guidelines. Movement Disorders, 27(3), 349–356.

Lucas-Jiménez, O., Ojeda, N., Peña, J., Díez-Cirarda, M., Cabrera-Zubizarreta, A., Gómez-Esteban, J. C., Gómez-Beldarrain, M. Á., & Ibarretxe-Bilbao, N. (2016). Altered functional connectivity in the default mode network is associated with cognitive impairment and brain anatomical changes in Parkinson’s disease. Parkinsonism & Related Disorders, 33, 58–64. 10.1016/j.parkreldis.2016.09.012

Lumme, V., Aalto, S., Ilonen, T., Någren, K., & Hietala, J. (2007). Dopamine D2/D3 receptor binding in the anterior cingulate cortex and executive functioning. Psychiatry Research: Neuroimaging, 156(1), 69–74. 10.1016/j.pscychresns.2006.12.012

Luo, C., Li, Q., Xia, Y., Lei, X., Xue, K., Yao, Z., Lai, Y., Martínez-Montes, E., Liao, W., Zhou, D., Valdes-Sosa, P. A., Gong, Q., & Yao, D. (2012). Resting state basal ganglia network in idiopathic generalized epilepsy. Human Brain Mapping, 33(6), 1279–1294. 10.1002/hbm.21286

Macleod, A. D., Taylor, K. S. M., & Counsell, C. E. (2014). Mortality in Parkinson’s disease: A systematic review and meta-analysis. Movement Disorders, 29(13), 1615–1622. 10.1002/mds.25898

Marek, K., Jennings, D., Lasch, S., Siderowf, A., Tanner, C., Simuni, T., Coffey, C., Kieburtz, K., Flagg, E., Chowdhury, S., Poewe, W., Mollenhauer, B., Klinik, P.-E., Sherer, T., Frasier, M., Meunier, C., Rudolph, A., Casaceli, C., Seibyl, J., … Taylor, P. (2011). The Parkinson Progression Marker Initiative (PPMI). Progress in Neurobiology, 95(4), 629–635. 10.1016/j.pneurobio.2011.09.005

Marek, S., & Dosenbach, N. U. F. (2018). The frontoparietal network: Function, electrophysiology, and importance of individual precision mapping. Dialogues in Clinical Neuroscience, 20(2), 133–140. 10.31887/DCNS.2018.20.2/smarek

Meireles, J., & Massano, J. (2012). Cognitive Impairment and Dementia in Parkinson’s Disease: Clinical Features, Diagnosis, and Management. Frontiers in Neurology, 3, 88. 10.3389/fneur.2012.00088

Menon, V. (2011). Large-scale brain networks and psychopathology: A unifying triple network model. Trends in Cognitive Sciences, 15(10), 483–506. 10.1016/j.tics.2011.08.003

Menon, V., & Uddin, L. Q. (2010). Saliency, switching, attention and control: A network model of insula function. Brain Structure and Function, 214(5), 655–667. 10.1007/s00429-010-0262-0

Nagano-Saito, A., Washimi, Y., Arahata, Y., Kachi, T., Lerch, J. P., Evans, A. C., Dagher, A., & Ito, K. (2005). Cerebral atrophy and its relation to cognitive impairment in Parkinson disease. Neurology, 64(2), 224–229. 10.1212/01.WNL.0000149510.41793.50

Narayanan, N. S., Cavanagh, J. F., Frank, M. J., & Laubach, M. (2013). Common medial frontal mechanisms of adaptive control in humans and rodents. Nature Neuroscience, 16(12), Article 12. 10.1038/nn.3549

Narayanan, Nandakumar & Albin, Roger. (2022). Cognition in Parkinson’s Disease—Google Books (1st ed., Vol. 269). Elsevier. https://books.google.com/books?hl=en&lr=&id=JwxSEAAAQBAJ&oi=fnd&pg=PP1&dq=cognition+in+parkinson%27s+disease+narayanan+albin&ots=P4imVA-wcE&sig=rzRZGj2XXmn4-Z1L4qspACEKk9Y#v=onepage&q=cognition%20in%20parkinson’s%20disease%20narayanan%20albin&f=false

Nasreddine, Z. S., Phillips, N. A., Bédirian, V., Charbonneau, S., Whitehead, V., Collin, I., Cummings, J. L., & Chertkow, H. (2005). The Montreal Cognitive Assessment, MoCA: A Brief Screening Tool For Mild Cognitive Impairment. Journal of the American Geriatrics Society, 53(4), 695–699. 10.1111/j.1532-5415.2005.53221.x

Obeso, J. A., Rodriguez-Oroz, M. C., Rodriguez, M., Lanciego, J. L., Artieda, J., Gonzalo, N., & Olanow, C. W. (2000). Pathophysiology of the basal ganglia in Parkinson’s disease. Trends in Neurosciences, 23, S8–S19. 10.1016/S1471-1931(00)00028-8

Parker, K. L., Chen, K.-H., Kingyon, J. R., Cavanagh, J. F., & Narayanan, N. S. (2015). Medial frontal _∼_4-Hz activity in humans and rodents is attenuated in PD patients and in rodents with cortical dopamine depletion. Journal of Neurophysiology, 114(2), 1310–1320. 10.1152/jn.00412.2015

Parker, K., Lamichhane, D., Caetano, M., & Narayanan, N. (2013). Executive dysfunction in Parkinson’s disease and timing deficits. Frontiers in Integrative Neuroscience, 7. https://www.frontiersin.org/articles/10.3389/fnint.2013.00075

Putcha, D., Ross, R. S., Cronin-Golomb, A., Janes, A. C., & Stern, C. E. (2015). Altered intrinsic functional coupling between core neurocognitive networks in Parkinson’s disease. NeuroImage: Clinical, 7, 449–455. 10.1016/j.nicl.2015.01.012

Putcha, D., Ross, R. S., Cronin-Golomb, A., Janes, A. C., & Stern, C. E. (2016). Salience and Default Mode Network Coupling Predicts Cognition in Aging and Parkinson’s Disease. Journal of the International Neuropsychological Society, 22(2), 205–215. 10.1017/S1355617715000892

Schölvinck, M. L., Maier, A., Ye, F. Q., Duyn, J. H., & Leopold, D. A. (2010). Neural basis of global resting-state fMRI activity. Proceedings of the National Academy of Sciences, 107(22), 10238–10243. 10.1073/pnas.0913110107

Seeley, W. W., Menon, V., Schatzberg, A. F., Keller, J., Glover, G. H., Kenna, H., Reiss, A. L., & Greicius, M. D. (2007a). Dissociable Intrinsic Connectivity Networks for Salience Processing and Executive Control. The Journal of Neuroscience, 27(9), 2349–2356.

Shafiei, G., Zeighami, Y., Clark, C. A., Coull, J. T., Nagano-Saito, A., Leyton, M., Dagher, A., & Mišić, B. (2019). Dopamine Signaling Modulates the Stability and Integration of Intrinsic Brain Networks. *Cerebral Cortex (New York*, NY), 29(1), 397–409. 10.1093/cercor/bhy264

Shima, A., Inano, R., Tabu, H., Okada, T., Nakamoto, Y., Takahashi, R., & Sawamoto, N. (2023). Altered functional connectivity associated with striatal dopamine depletion in Parkinson’s disease. Cerebral Cortex Communications, 4(1), tgad004. 10.1093/texcom/tgad004

Singh, A., Cole, R. C., Espinoza, A. I., Evans, A., Cao, S., Cavanagh, J. F., & Narayanan, N. S. (2021). Timing variability and midfrontal ∼4 Hz rhythms correlate with cognition in Parkinson’s disease. Npj Parkinson’s Disease, 7(1), Article 1. 10.1038/s41531-021-00158-x

Singh, A., Cole, R. C., Espinoza, A. I., Wessel, J. R., Cavanagh, J. F., & Narayanan, N. S. (2023). Evoked mid-frontal activity predicts cognitive dysfunction in Parkinson’s disease. *Journal of Neurology*, Neurosurgery & Psychiatry, jnnp-2022–330154. 10.1136/jnnp-2022-330154

Singh, A., Richardson, S. P., Narayanan, N., & Cavanagh, J. F. (2018). Mid-frontal theta activity is diminished during cognitive control in Parkinson’s disease. Neuropsychologia, 117, 113–122. 10.1016/j.neuropsychologia.2018.05.020

Sridharan, D., Levitin, D. J., & Menon, V. (2008). A critical role for the right fronto-insular cortex in switching between central-executive and default-mode networks. Proceedings of the National Academy of Sciences of the United States of America, 105(34), 12569– 12574. 10.1073/pnas.0800005105

Summerfield, C., Junqué, C., Tolosa, E., Salgado-Pineda, P., Gómez-Ansón, B., Martí, M. J., Pastor, P., Ramírez-Ruíz, B., & Mercader, J. (2005). Structural Brain Changes in Parkinson Disease With Dementia: A Voxel-Based Morphometry Study. Archives of Neurology, 62(2), 281–285. 10.1001/archneur.62.2.281

Tessitore, A., Cirillo, M., & De Micco, R. (2019). Functional Connectivity Signatures of Parkinson’s Disease. Journal of Parkinson’s Disease, 9(4), 637–652. 10.3233/JPD-191592

Tessitore, A., Esposito, F., Vitale, C., Santangelo, G., Amboni, M., Russo, A., Corbo, D., Cirillo, G., Barone, P., & Tedeschi, G. (2012). Default-mode network connectivity in cognitively unimpaired patients with Parkinson disease. Neurology, 79(23), 2226–2232. 10.1212/WNL.0b013e31827689d6

Tikàsz, A., Potvin, S., Dugré, J. R., Fahim, C., Zaharieva, V., Lipp, O., Mendrek, A., & Dumais, A. (2020). Violent Behavior Is Associated With Emotion Salience Network Dysconnectivity in Schizophrenia. Frontiers in Psychiatry, 11. https://www.frontiersin.org/articles/10.3389/fpsyt.2020.00143

Uc, E., Anjum, F., Dasgupta, S., & Narayanan, N. (2023). Resting-state EEG Predicts Cognitive Impairment in Parkinson’s Disease (P6-11.015). Neurology, 100(17 Supplement 2). 10.1212/WNL.0000000000203685

Uddin, L. Q. (2016). Salience Network of the Human Brain. Academic Press.

Ueno, D., Matsuoka, T., Kato, Y., Ayani, N., Maeda, S., Takeda, M., & Narumoto, J. (2020). Individual Differences in Interoceptive Accuracy Are Correlated With Salience Network Connectivity in Older Adults. Frontiers in Aging Neuroscience, 12. https://www.frontiersin.org/articles/10.3389/fnagi.2020.592002

Vogel, J. W., Corriveau-Lecavalier, N., Franzmeier, N., Pereira, J. B., Brown, J. A., Maass, A., Botha, H., Seeley, W. W., Bassett, D. S., Jones, D. T., & Ewers, M. (2023). Connectome-based modelling of neurodegenerative diseases: Towards precision medicine and mechanistic insight. Nature Reviews Neuroscience, 1–20. 10.1038/s41583-023-00731-8

Vogt, B. A. (2019). Chapter 13—Cingulate cortex in Parkinson’s disease. In B. A. Vogt (Ed.), Handbook of Clinical Neurology (Vol. 166, pp. 253–266). Elsevier. 10.1016/B978-0-444-64196-0.00013-3

Webb, C. A., Israel, E. S., Belleau, E., Appleman, L., Forbes, E. E., & Pizzagalli, D. A. (2021). Mind-Wandering in Adolescents Predicts Worse Affect and Is Linked to Aberrant Default Mode Network-Salience Network Connectivity. Journal of the American Academy of Child and Adolescent Psychiatry, 60(3), 377–387. 10.1016/j.jaac.2020.03.010

Whitfield-Gabrieli, S., & Nieto-Castanon, A. (2012). Conn: A Functional Connectivity Toolbox for Correlated and Anticorrelated Brain Networks. Brain Connectivity, 2, 125–141. 10.1089/brain.2012.0073

Zadikoff, C., Fox, S. H., Tang-Wai, D. F., Thomsen, T., de Bie, R. M. A., Wadia, P., Miyasaki, J., Duff-Canning, S., Lang, A. E., & Marras, C. (2008). A comparison of the mini mental state exam to the montreal cognitive assessment in identifying cognitive deficits in Parkinson’s disease. Movement Disorders, 23(2), 297–299. 10.1002/mds.21837

Zgaljardic, D. J., Borod, J. C., Foldi, N. S., & Mattis, P. (2003). A Review of the Cognitive and Behavioral Sequelae of Parkinson’s Disease: Relationship to Frontostriatal Circuitry. Cognitive and Behavioral Neurology, 16(4), 193.

